# A Multimodal Assessment of the Heterogeneous Nature of Post-mTBI Dizziness

**DOI:** 10.1101/2022.07.12.22277563

**Authors:** Ryan Pelo, Elle Gaudette, Leah Millsap, Cecilia Martindale, Leland E. Dibble, Melissa M. Cortez, Peter C. Fino

## Abstract

Dizziness after mild traumatic brain injury (mTBI) is commonly attributed to impairment within the vestibular system. However, oculomotor, mobility, and autonomic dysfunction can also contribute to patient-reported dizziness. The purpose of this preliminary study was to examine whether a multimodal battery of assessments could help explain patient-reported dizziness after mTBI. Twenty-three participants with concussion-related symptoms completed the Dizziness Handicap Inventory (DHI) to evaluate burden imposed by dizziness on daily activities and a battery of tests designed to incorporate domains that have been shown to contribute to dizziness (e.g., vestibular, oculomotor, balance and mobility, and autonomic dysfunction). Specific outcomes included quantitative variables obtained from: Vestibular Ocular Motor Screening (VOMS); standing for 30 seconds with feet together, eyes closed, with hands on their hips on both firm and foam surfaces; walking for one minute at a comfortable pace; and a Head-up Tilt (HUT) Test. Univariate associations between DHI and individual measures were assessed, and a backwards-stepwise regression model determined the multi-variable association. There were no strong associations and only a few moderate associations between individual functional measure and DHI total score. A total of eight variables had univariate correlation coefficients larger than 0.20 in magnitude. The final model from the backwards-stepwise procedure explained 69% of the variance in DHI and retained only three variables: peak turning speed from the one-minute walk; mean blood pressure (MBP) during the HUT; and the total VOMS score. Isolated assessments of individual domains of function have weak-to-moderate associations with post-mTBI dizziness. Conversely, a multivariable model that contained measures of mobility, autonomic function, and symptomatic complaints to vestibular and ocular provocation explained 69% of the variance in dizziness. These results suggest that dizziness is physiologically heterogeneous in nature and support the use of multi-domain assessments in patients with dizziness after mTBI.

## INTRODUCTION

Mild traumatic brain injuries (mTBIs) affect millions of people each year and cause symptoms and functional deficits that can have persistent effects.^1,2^ Dizziness is the second most common symptom following mTBI and is reported by nearly 80% of individuals immediately after injury.^3,4^ While dizziness can sometimes resolve spontaneously, over 25% of individuals experience symptoms of dizziness over one year after their mTBI.^5^ This dizziness can lead to limitations in daily life activities and decreased quality of life,^6^ making dizziness an important symptom to assess and treat in the rehabilitation of patients with mTBI.

Assessments of dizziness after mTBI often rely on patient self-report using symptom checklists. However, single-item responses that ask patients to rate overall dizziness are nonspecific and are thus not recommended as standalone assessment measures.^7–10^ The Dizziness Handicap Inventory (DHI) was developed as a structured self-report assessment of situation-dependent impact of dizziness symptoms,^11^ and is recommended over symptom severity ratings.^7,12^ Yet, while the DHI provides insight into the daily impact of dizziness, it does not yield information on the underlying cause.

Despite common treatment models assigning dizziness symptoms to a ‘vestibular’-based symptom subtype / cluster,^13–16^ self-reported disability due to dizziness cannot be easily attributed to a single system; vestibular, oculomotor, balance and mobility, and autonomic dysfunction can all be independent contributors to dizziness.^7,17–23^ The vestibular system functions to sense movement and orientation of the head and body in space, which, if altered, can produce symptoms of dizziness.^13,24^ Within the vestibular domain, a recent study found no clear association between patient-reported dizziness or imbalance and peripheral vestibular dysfunction, suggesting deficits in central sensorimotor integration are more prevalent after mTBI than peripheral vestibular disorders.^25^ Dizziness within the ocular-motor domain often stems from dysfunction in the ability to maintain gaze stabilization either with head movement or when the head is still and there is movement within the visual field.^26^ Although often working in conjunction with the vestibular system and at times providing complementary information, the ocular motor system is critical in understanding space and the environment surrounding the body.^27^ Balance and mobility, such as turning kinematics, gait speed, and postural sway, involve more complex integration of peripheral sensory signals that may indicate central sensory integration problems, rather than peripheral sensory dysfunction and have all been found to be altered after mTBI.^28–31^ Further, speculation suggests that altered movement behaviors may be self-selected in an effort to reduce symptom exacerbation, including limiting dizziness by reducing rapid rotations of the head and body.^32^ Finally, autonomic nervous system control of blood pressure (BP) and heart rate (HR) in response to positional changes is critical in the maintenance of consistent cerebral blood flow.^33,34^ An inability to maintain cerebral blood flow by modulating BP and HR based on situational demands can contribute to dizziness, syncope, and other symptoms that overlap with dizziness such as light-headedness.^35,36^

As dizziness can be linked to multiple domains, domain-directed functional assessments can provide localizing information about the origin of the dizziness and guide clinical treatment. For example, orthostatically induced dizziness (or lightheadedness) may reflect measurable physiological changes in cardiovascular function that resolve in parallel with post-mTBI symptom burden.^37^ Yet, most functional assessments have poor-to-moderate associations with dizziness.^38^ The Vestibular Ocular Motor Screen (VOMS), a common clinical screen used to assess vestibular and ocular motor impairments after mTBI,^39^ has only moderate association with overall self-reported disability due to dizziness, as assessed through the DHI Total score.^40^ The lack of strong relationship between functional assessment and dizziness may arise from trying to associate uni-dimensional assessments of function to the heterogeneous concept of dizziness. While the VOMS does associate with selected vestibular-based items within the DHI, using sub-scores of the DHI is not recommended and offers only a limited view of the patient’s experience with dizziness.^41^ A more complete understanding of the scope and associated physiological changes related to dizziness after mTBI is needed.

The purpose of this study was to provide a preliminary evaluation of the origins of dizziness after mTBI using a multimodal battery of functional assessments. We hypothesized that greater levels of self-reported disability due to dizziness after mTBI would be associated with increased impairment within each isolated domain (e.g., including vestibular, oculomotor, balance and mobility, and autonomic dysfunction), but that dizziness would be best explained by a combination of factors that span the multiple mechanistic domains of dizziness.

## MATERIALS and METHODS

### Participants

Twenty-eight participants gave informed written consent and participated in a collection of broader IRB-approved studies on mTBI. Inclusion criteria for this analysis was: between the ages of 18-50 years and diagnosed with an mTBI by a physician within the past year; mTBI diagnosis was confirmed at enrollment using Veterans Affairs/Department of Defense mTBI diagnostic criteria^42^; all subjects remained symptomatic at the time of screening, and reported that they were currently experiencing at least one symptom of mTBI, assessed through the Neurobehavioral Symptom Inventory (NSI).

Exclusion Criteria: Under the age of 18 or age 51 or older, diagnosis of any other injury, medical, or neurological illness that could potentially explain balance deficits (e.g., central or peripheral nervous system disease, stroke, greater than mTBI, lower extremity amputation, recent lower extremity or spine orthopedic injury); met criteria for moderate to severe substance-use disorder within the past month as defined by DSM-V; pregnancy (balance considerations); and/or displayed behavior that would significantly interfere with the validity of data collection or safety during the study.

### Dizziness Assessment

At the start of the session, participants completed the DHI to evaluate burden imposed by dizziness.^11^ The DHI is a 25-item self-report measure that assesses the self-perceived handicap one experiences during everyday life due to dizziness.^11^ Items are scored as no = 0, sometimes = 2, and yes = 4, and then summed with a range of 0-100.^11^ Items can further be broken down into three domains: functional, emotional, and physical.^11^ The DHI has shown high test-retest reliability with both the total score and the sub-domains, and has shown to be valid with a brain injury population.^11,43,44^

### Vestibular and Ocular Motor Screening

Participants also completed the Vestibular Ocular Motor Screen (VOMS), a tool designed to measure symptomatic changes before and after vestibular-ocular tasks in people with mTBI.^39^ The screening tests for symptom provocation during seven total categories: Smooth Pursuits, Saccades (vertical and horizontal), Convergence, Vestibular-Ocular Reflexes (vertical and horizontal), and Visual Motion Sensitivity. Symptoms of nausea, fogginess, dizziness, and headache were reported on a point scale of 0 (no symptoms) to 10 (highest symptom rating) at baseline, prior to beginning screening and after each task. The change in symptoms was determined by comparing the self-reported symptom severity to the baseline symptoms; any increase in symptoms indicated a positive test item.^39^ The number of items with at least a one point change (i.e., positive item) was used as the total VOMS score, ranging from 0 (no reported symptom changes in all seven categories) to 7 (reported symptom changes in all seven categories), to include both vertical and horizontal testing of Saccades and Vestibular-Ocular Reflexes.

### Balance and Mobility Performance

To assess balance and mobility, participants completed two standing balance tasks and one walking task. The balance tasks consisted of standing quietly for 30 seconds with feet together, eyes closed, and hands on their hips on both firm and foam surfaces. Additionally, participants completed a walking task that consisted of walking between two lines 6 m apart at their preferred walking speed for one minute. Participants were asked to walk at their normal, comfortable walking speed past the 6 m marking line, turn around after passing the line, walk back towards the starting line, and continue walking back and forth between these lines for one minute. For both tasks, objective kinematic data was recorded using wearable inertial measurement units (IMUs; Opals v2, APDM Inc., Portland, OR) that were placed on the participants’ feet, lumbar spine (approximately L3-L5 region), sternum, and forehead. The IMUs sampled tri-axial acceleration and angular velocity data at 128 Hz. Standard commercial software (Moveo, APDM Inc, Portland OR) was used to compute the root-mean-square (RMS) of lumbar acceleration in the mediolateral direction during the quiet standing tasks, gait speed during the one-minute walk task, and peak turning speed during the turns of the one-minute walk task. ^45,46^

### Physiological Function

To assess cardiovascular autonomic function, participants completed a 10-minute head-up tilt (HUT) table test. Participants were secured to the tilt table with their feet flush to a footboard to minimize movement and potential artifact during the tilt; blood pressure and heart rate were assessed using a non-invasive beat-to-beat blood pressure device (Nexfin Model 2, Edwards Lifesciences/BMEYE, Amsterdam, Netherlands; and CNAP Monitor 500, CNSystems Medizintechnik GmbH, Graz, Austria). ^47–50^ A sling was used to maintain their upper extremity at heart level for the duration of testing. An external brachial blood pressure was also used to verify the accuracy of continuous measurements at regular intervals throughout the testing, coincident with symptom checks. Testing was performed in a dimly lit room, where relaxation was promoted and distractions were minimized (e.g., participants were instructed to avoid speaking unless necessary, such as in the case of intolerable symptoms per participant judgment). Data was considered based on three pre-defined segments: the initial five-minute rest period (Supine); ten minutes of HUT in which the head of the table was elevated to 70° with respect to the horizontal; and the final five minutes in which the tilt table was returned to a supine position (Return-to-Supine). Symptoms were periodically monitored throughout the duration of the test to ensure patient safety and wellbeing but were not considered in the analysis. Blood pressure and heart rate data were captured using Testworks software (WR Medical Electronics Co., Stillwater, MN) and exported for offline analysis of mean blood pressure (MBP), mean heart rate (MHR), blood pressure variability (BPCV), and heart rate variability (HRCV) during each segment. The change in MBP, MHR, BPCV, and HRCV between supine and HUT segments was also calculated and used as an outcome variable.

### Statistical Analysis

Pearson correlation coefficients assessed the univariate relationship between each variable and DHI total scores. Confidence intervals for each correlation coefficient were obtained using a bootstrapping procedure with 100 iterations. Correlation strength was interpreted as ρ ≤ 0.35 indicating weak, 0.35 < ρ ≤ 0.67 indicating moderate, and ρ > 0.67 indicating strong associations.^51^ To determine if a sparse combination of individual items could better explain variance in post-mTBI DHI, a multi-step process was implemented. To reduce the dimensionality of the variables, only variables with a correlation coefficient greater than 0.20 in magnitude were retained for future multivariable modeling. Next, a backwards-stepwise linear regression model was implemented to determine the combination of variables that associated with the DHI total score, with the change in Bayesian Information Criterion (BIC) used to determine the removal of each term and the final model. A total of 100 bootstrapped linear models were then fit using the final combination of variables as independent predictors of the DHI total score, with the robustness of the model fit assessed using the mean and 95% confidence intervals of the bootstrapped models’ R^2^ values. All statistical analyses were performed in MATLAB R2020a and the Statistics and Machine Learning Toolbox (The Mathworks, Inc., Natick, MA, USA).

## RESULTS

### Sample Characteristics

A total of 22 participants mean (SD) age = 32.2 (8.1) years completed all components of the study and were included in this analysis; data from five participants were incomplete due to equipment malfunctions during in the data collection, and one participant was excluded for being greater the one year post-injury. The mean DHI score for the 22 included participants was 30.0 (SD = 27.6) and mean NSI score was 29.7 (SD = 20.6). Time since injury ranged from 3-267 days post mTBI (median 9.5 days) (Table 1).

**TABLE 1.**
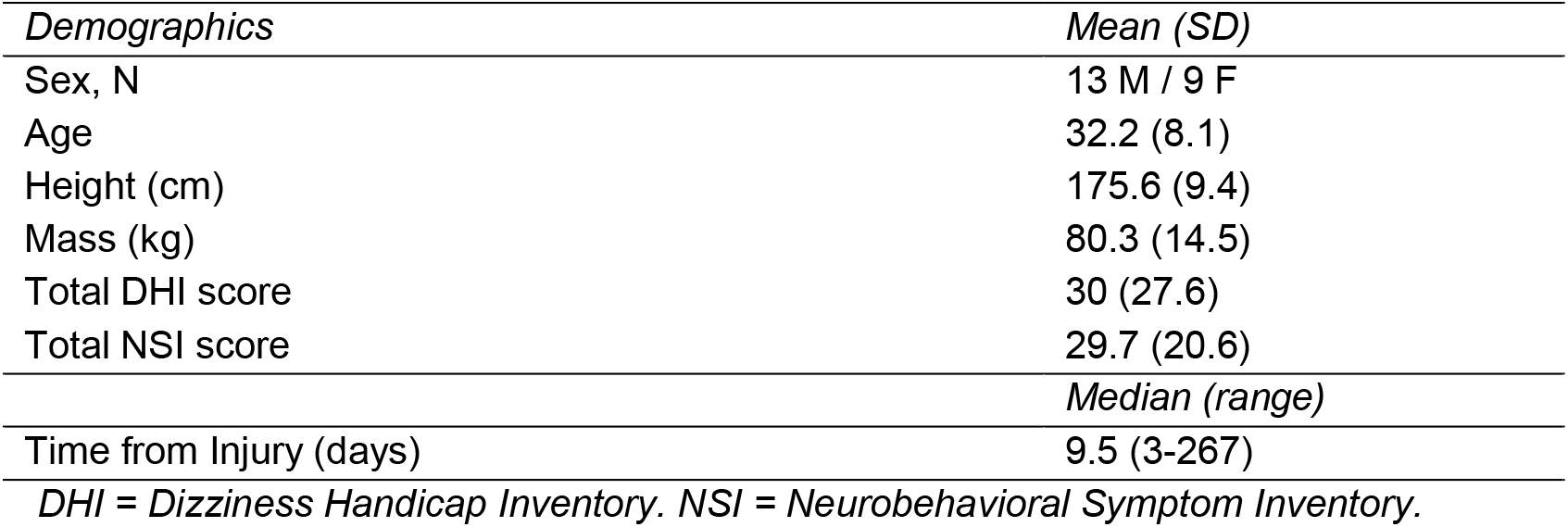
Descriptive statistics for overall sample.

| *Demographics* | *Mean (SD)* |
| --- | --- |
| Sex, N | 13 M / 9 F |
| Age | 32.2 (8.1) |
| Height (cm) | 175.6 (9.4) |
| Mass (kg) | 80.3 (14.5) |
| Total DHI score | 30 (27.6) |
| Total NSI score | 29.7 (20.6) |
| *Median (range)* |  |
| Time from Injury (days) | 9.5 (3-267) |

*DHI = Dizziness Handicap Inventory. NSI = Neurobehavioral Symptom Inventory.*

### Correlations of DHI and Individual Measurements

There were no strong associations and only a few moderate associations between individual functional measure and DHI total score (Table 2). Amongst all measures, the VOMS total score was most associated with DHI with a moderate association (Pearson’s ρ = 0.49).

**TABLE 2.** Correlation and descriptive statistics of all measurements and DHI. Bolded variables were those that met the cutoff (*ρ* > 0.20) for inclusion into the multivariable regression model.

| *Motor Measurements* | *Mean (SD)* | *Pearson Correlation Coefficient,*<br>*$\rho$ (95% CI)* |
| --- | --- | --- |
| **Turning Velocity (deg/s)** | **190.0 (26.3)** | **-0.27 (-0.31, -0.23)** |
| Mean Gait Speed (m/s) | 1.18 (0.14) | -0.15 (-0.23, -0.12) |
| ML RMS Sway, Firm (m/s<sup>2</sup>) | 0.09 (0.04) | 0.15 (0.14, 0.24) |
| ML RMS Sway, Foam (m/s<sup>2</sup>) | 0.22 (0.11) | -0.15 (-0.18, -0.09) |
| *Physiological Measurements* |  |  |
| MBP Supine (mmHg) | 85.0 (10.2) | 0.08 (0.06, 0.13) |
| BP CV Supine (%) | 5.8 (2.3) | 0.16 (0.11, 0.19) |
| MHR Supine (bpm) | 63.0 (11.5) | 0.16 (0.14, 0.22) |
| HR CV Supine (%) | 6.5 (3.3) | 0.02 (-0.06, -0.04) |
| **MBP HUT (mmHg)** | **92.6 (12.2)** | **0.36 (0.33, 0.40)** |
| **BP CV HUT (%)** | **7.5 (4.5)** | **0.20 (0.14, 0.25)** |
| MHR HUT (bpm) | 81.9 (11.5) | 0.14 (0.12, 0.20) |
| **HR CV HUT (%)** | **8.8 (4.8)** | **0.45 (0.39, 0.49)** |
| **Change in MBP (mmHg)** | **7.6 (11.5)** | **0.23 (0.17, 0.28)** |
| Change in MHR (bpm) | 19.0 (6.6) | -0.04 (-0.11, -0.01) |
| **Change in BP CV (%)** | **1.7 (4.0)** | **0.25 (0.18, 0.29)** |
| **Change in HR CV (%)** | **-0.3 (10.9)** | **-0.21 (-0.30, -0.13)** |
| *Vestibular-Ocular Measurements* |  |  |
| **VOMS Total Score** | **4.1 (2.5)** | **0.49 (0.47, 0.54)** |

*ML RMS = Mediolateral Root Mean Squared, MBP = Mean Blood Pressure, MHR = Mean Heart Rate, BPCV = Blood Pressure Variability, HRCV = Heart Rate Variability, HUT = Head-Up Tilt, VOMS = Vestibular Ocular Motor Screening*
*Bold measures were retained for multivariable analysis*

### Backwards-Stepwise Regression Analysis

A total of eight variables had univariate correlation coefficients larger than 0.20 in magnitude and were entered into the backwards-stepwise linear regression model. The final model retained only three variables: peak turning speed from the one-minute walk; MBP during the HUT; and the total VOMS score (Table 3). The multivariable model containing these three variables explained 69.2% of the variance in DHI scores (Figure 1). The bootstrapped models yielded R^2^ values ranging from 0.43 to 0.90, with a mean (95% confidence interval) R^2^ of 0.70 (0.68, 0.72).

**TABLE 3.** Final Model Summary Output from the Backwards Stepwise Regression.

| *Regression Statistics* |  |
| --- | --- |
| Number of Observations | 22 |
| Error Degrees of Freedom | 18 |
| Root Mean Squared Error | 16.5 |
| R-Squared | 0.692 |
| Adjusted R-Squared | 0.641 |
| F-Statistic vs Constant Model | 13.5 |
| P-value | 0.00007 |

*Estimated Coefficients*
|  | *Estimate* | *Standard Error* | *P Value* |
| --- | --- | --- | --- |
| Intercept | 22.9 | 41.4 | 0.58 |
| Average Peak Turning Speed (deg/s) | -0.60 | 0.15 | 0.001 |
| MBP HUT (mmHg) | 0.91 | 0.30 | 0.007 |
| VOMS Total Scores (positive items) | 9.0 | 1.6 | < 0.001 |

*MBP = Mean Blood Pressure, HUT = Head-Up Tilt, VOMS = Vestibular Ocular Motor Screening*

**Figure 1.**
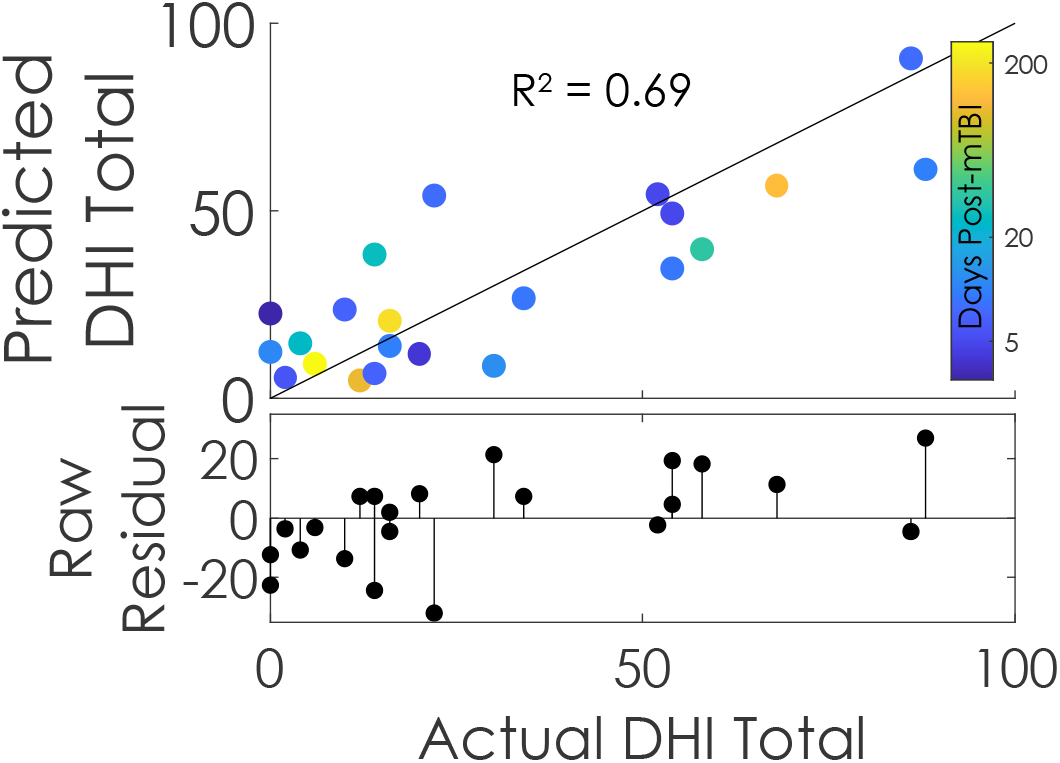
Final output of the backwards-stepwise regression model containing the variables: peak turning speed; MBP during HUT; and Total VOMS score. Predicted DHI Total = β0 + β1* MBP during HUT + β2* Total VOMS score + β3* peak turning speed.

## DISCUSSION

The results of this study found weak and inconsistent associations between DHI score and isolated assessments of individual domains (e.g., including vestibular, oculomotor, balance and mobility, and autonomic dysfunction), whereas a multivariable model that contained measures of mobility, autonomic function, and symptomatic complaints to vestibular and ocular provocation explained 69% of the variance in DHI score. This finding supports the principle that self-reported disability due to dizziness is physiologically heterogeneous in nature and is best explained through consideration of a combination of vestibular, oculomotor, balance and mobility, and autonomic function assessments. Increased awareness of the heterogeneous nature of dizziness and its multiple contributing domains is a significant step toward an improved mechanistic understanding of the multivariate nature of dizziness following mTBI. While preliminary given the small sample size, these results present the first sample of mTBI participants with objective quantification of autonomic function, balance and mobility, and symptomatic provocation from the VOMS, and lay a foundation for future studies to incorporate multi-modal assessments that consider each of these domains.

### A Multimodal Clinical Assessment of Dizziness Provides Insight into Affected Domains

While we are not aware of any studies that have comprehensively assessed vestibular, oculomotor, balance and mobility, and autonomic function in relation to dizziness after mTBI, our results align with the recommendations put forth by the 2020 Clinical Practice Guidelines for Physical Therapy after concussion / mTBI.^52^ These guidelines indicate that the combined assessment of vestibulo-oculomotor, autonomic (e.g., exertional tolerance), motor, and cervical musculoskeletal function is warranted for patients with self-reported dizziness. These guidelines are generated through systematic review but have been lacking empirical evidence to date. Our results provide support for this approach, favoring a multimodal battery of assessments, rather than singular subtyping of symptoms. The present result – that dizziness is multi-dimensional – is also at odds with current subtype classification guidelines that drive current rehabilitation practices.^13–16^ For example, the assignment of a person’s dizziness to a ‘vestibular’-based symptom subtype / cluster may ultimately mislabel a significant portion of the symptom-related disability, leading the rehabilitation provider to neglect directed treatment of potential contributing factors and / or physiological domains. Even the VOMS Total Score, which is routinely used to triage patients as the ‘vestibular’-based subtype, encompasses multiple physiological/mechanistic domains within the assessment.

### Multiple Mechanisms Contribute to the Heterogeneous Nature of Post-mTBI Dizziness

The final multivariable model that explained over 69% of the variance in DHI symptoms included variables representing three distinct physiological domains that provide complementary information about dizziness: symptom provocation from vestibular and ocular tasks (VOMS Total Score), complex mobility (turning speed), and cardiovascular autonomic function (mean BP during HUT). Considering the VOMS Total Score is a direct measurement of symptoms; it is unsurprising that this variable is important to explain a patient’s self-perceived dizziness handicap. However, the VOMS provides little information about the underlying vestibular or ocular impairment.^39,53^ Further assessments of function are needed to determine *how* these tasks elicit symptoms; the vestibular and ocular motor systems work in conjunction with each other and at times provide overlapping information that may conflict to elicit symptoms.^27,54^ Further, the large percentage of dizziness that is *not* explained by the VOMS – the VOMS alone explained only 24% of the variance in DHI – suggests that functional activities common to daily living may be important to self-reported dizziness handicap.

The inclusion of peak turning speed in our multivariable model indicates that complex mobility tasks that are common to daily life are an important feature to consider in a patient’s reported dizziness. The peak turning speed measured here was obtained as participants were instructed to walk at their self-selected pace – thus, differences in gait or turning speed could indicate *either* a central sensory integration problem *or* a conscious self-monitoring of movement to avoid exacerbating symptoms. Previous speculation suggested individuals may restrict the speed or amplitude of movements in an effort to reduce dizziness.^32,55^ Previous studies in a centrifuge have found that the cerebrovascular regulatory response to fast rotations is also a contributing factor in the development of dizziness and nausea in healthy adults, suggesting that slower or abnormal self-selected mobility after mTBI may be – at least in part - a compensatory strategy for impaired cerebrovascular control.^35,56^

These results provide further support for more directed assessment of cardiovascular autonomic function after mTBI, particularly in patients with reported dizziness. Cardiovascular autonomic function assessed through the HUT test – as done here – predominantly evaluates baroreceptor-mediated sympathetic function through an orthostatic stress.^57^ During the test, a shift in blood volume into the lower extremities should result in unloading of the cardiopulmonary and arterial baroreceptors which triggers vasoconstriction and increased HR in order to maintain appropriate cerebral blood flow.^58,59^ The retention of MBP during the HUT in our final multivariable model supports prior studies suggesting abnormal sympathetic augmentation upon baroreceptor activation in people with post-mTBI dizziness.^60–62^ Given that prior studies have reported variable patterns of altered HR and BP control following mTBI,^34,63,64^ and the preliminary nature of this study, future studies including HR and BP measures of cardiovascular autonomic function are needed.^37^ In the meantime, these data support clinical (bedside) assessment of standard orthostatic vital signs^37,65^, which may provide an informative physiological correlate to symptoms of dizziness, provide an additional objective parameter to follow during rehabilitation, and direct treatment approaches.^66^

## Limitations

Our relatively small sample size and heterogeneous study group (representing a wide range of ages, mechanisms of injury, and time since mTBI) limit the generalizability of our findings, and merit follow-up study. While it is possible that age- or recovery-duration may influence the association between each variable and DHI score, our results appear robust to variations in time since injury (see color bar on Figure 1). However, we expect that the effect of time since injury would differ if both symptomatic and asymptomatic individuals were included. The wide timeline of testing post-mTBI (3-267 days) likely added heterogeneity, but also suggests these findings may generalize beyond a specific window of recovery (e.g., acute) to encompass any individual with persisting dizziness after mTBI. Finally, the assessments performed within our study were selected based on their representation of each domain, but do not necessarily represent a complete mechanistic understanding of the pathophysiology present within each domain. Thus, future larger, longitudinal studies might further explore the relationship of age, timing of injury, and/or recovery on the factors contributing to dizziness related disability.

## CONCLUSION

The heterogeneous nature of dizziness lends itself to the need for a multimodal assessment to better understand the origin of impairment after mTBI. A singular ‘vestibular’-based symptom subtype can limit the mechanistic understanding of dizziness and may contribute to inapt or non-specific treatments. Our results suggest that a combination of domains that includes vestibular, oculomotor, balance and mobility, and autonomic assessments provide a more complete understanding of the physiological origins of dizziness, providing important opportunities for more mechanism-targeted treatment interventions.

## Data Availability

All data produced in the present study will be available through the Federal Interagency Traumatic Brain Injury Research (FITBIR) Informatics System.

## ACKNOWLEDGEMENTS

The authors would like to acknowledge Sarah Hill, Shu Yang, and Ben Cassidy for their assistance in collecting the data.

## AUTHORSHIP CONFIRMATION/CONTRIBUTION STATEMENT

**Ryan Pelo:** Writing – original draft (lead); data curation (equal); writing – review and editing (equal). **Elle Gaudette:** Writing – original draft (supporting); data curation (equal); formal analysis (supporting); writing – review and editing (equal). **Leah Millsap:** Data curation (equal); writing – review and editing (equal), **Cecilia Martindale:** Data curation (equal); writing – review and editing (equal), **Leland E. Dibble:** Writing – review and editing (equal) **Melissa M. Cortez:** Conceptualization (supporting); formal analysis (supporting); writing – review and editing (equal). **Peter C. Fino:** Conceptualization (lead); formal analysis (lead); writing – original draft (supporting); writing – review and editing (equal)

